# Critical temperature thresholds for identifying vulnerability to heat-related excess cardiovascular morbidity and mortality

**DOI:** 10.1101/2025.08.28.25334692

**Authors:** Peter M. Graffy, Benjamin W. Barrett, Daniel E. Horton, Abel N. Kho, Norrina B. Allen

## Abstract

**Background:** Extreme heat is a known environmental hazard linked to increased cardiovascular disease (CVD) morbidity and mortality, yet most studies fail to evaluate both acute and long-term effects. We assessed heat-related CVD, and its subtypes: coronary heart disease (CHD), myocardial infarction (MI), and stroke, morbidity and mortality across multiple temporal scales, identifying critical temperature thresholds for short- and long-term exposure.

**Methods:** We analyzed neighborhood-level death and emergency department (ED) visit records for CVD in Chicago from 2011–2022, linked to sociodemographic characteristics and temperature and humidity estimates. Using generalized additive models with smooth splines, we estimated excess heat-related rates across daily, monthly, and annual temporal scales. We applied K-means clustering and principal component analysis to classify Chicago community areas by heat vulnerability.

**Results:** Higher temperatures were significantly associated with increased cardiovascular mortality across all temporal scales, but not morbidity. Daily maximum temperatures ≥101.3°F were linked to 0.098 excess CHD deaths per 100,000 (95% confidence interval [CI]: 0.092–0.104, p=0.037), while a 3-day average ≥100.4°F corresponded to 0.109 excess CVD deaths per 100,000 (95% CI: 0.084–0.139, p *<* 0.001), which may lead to ≈3 daily CVD deaths in Chicago are heat-related during days above 100°F. Monthly associations were also seen for CHD mortality (3.30 excess deaths per 100,000, [95% CI: 2.91–3.66], p=0.001). No significant associations were found for MI or stroke. Mortality clustering revealed four distinct vulnerability profiles, with variation linked to socioeconomic and racial/ethnic characteristics.

**Conclusions:** We identified critical temperature thresholds associated with cardiovascular mortality across temporal and spatial scales. Findings support targeted public health interventions, including heat warning systems and adaptive strategies, tailored to community vulnerability.

## INTRODUCTION

Cardiovascular disease (CVD) is the leading cause of morbidity and mortality in the United States (US) and heat exposure increases the burden of disease. After initial declines in CVD mortality over the last decade, the age-adjusted mortality rate climbed 9.3% to nearly 500 per 100,000 by 2022 [1, 2]. CVD event incidence is impacted by heat exposure, which is expected to increase in severity and duration because of climate change [3, 4]. Excess cardiovascular deaths due to extreme heat are estimated to increase 161-232% by 2065 in the US [5]. Urban areas with a large heat island effect are the most vulnerable to heat-related CVD, with Chicago projected to have the largest burden of annual CVD morbidity and mortality of all major US cities by 2090 [6–8]. Identifying the areas of Chicago that are most susceptible to heat-related CVD will be important for resource allocation and preventive efforts in the face of consistently higher-than-average temperatures.

There are no standard definitions for extreme heat or heatwaves and little work has been done to concomitantly assess both acute and long-term impacts of heat on communities [9]. Many studies have characterized the impact of heat exposure on CVD hospitalizations and mortality [10–12]. However, these studies are often unable to identify vulnerable areas and microclimates since they are generally at a large geographic level, use low resolution temperature estimate methods, or fail to capture the effects of both short- and long-term heat exposure on health outcomes. Prior studies have estimated the relative risk of CVD using predefined percentile thresholds for temperature which provide subjective associations that potentially miss larger patterns [12, 13]. Likewise, the pathophysiological mechanisms of CVD in the presence of heat exposure manifest differently depending on CVD subtype, necessitating a spectrum of temperature thresholds to determine risk. In this study, we evaluated the impact of temperature during warm months on excess CVD emergency department (ED) visits and mortality rates within Chicago community areas for various temporal scales. The purpose of this study was to isolate the effect of cumulative heat exposure on CVD morbidity and mortality using high resolution temperature estimates, identify population-based temperature thresholds for CVD and its subtypes, and characterize community-level vulnerability to short- and long-term heat exposure.

## MATERIALS AND METHODS

This was an IRB-approved retrospective ecological time series study performed in collaboration with seven academic medical centers in Chicago and the Illinois and Chicago Departments of Public Health. We included decedents and ED patients age ≥ 18 years with an address that spatially corresponded to a Chicago community area between 2011 and 2022 for months May through September.

Chicago has 77 community areas which were originally established in 1920 and whose boundaries have remained unchanged since 1980. The most recent geographic shapefile for the community areas is publicly available through the City of Chicago Data Portal (https://data.cityofchicago.org/). Annual sociodemographic estimates of community area characteristics were queried from the American Community Survey (ACS) 5-year tables. The variables selected include annual estimates of total population, education level, racial composition, sex distribution, employment status, median income, and median age for each community area across the entire 2011-2022 study period.

Daymet is a publicly available product developed by Oak Ridge National Laboratory which contains daily estimates of maximum temperature (T_max_), minimum temperature (Tmin), and vapor pressure (VP) at a 1 km2 spatial resolution across North America. Daily mean estimates of T_max_, Tmin, and VP were aggregated for each community area for the duration of the study period by spatially joining the community area shapefile with a raster brick of Daymet estimates and calculating the weighted mean of Daymet grid cells within each respective community area. Daily mean temperature (Tmean) and relative humidity (RH) values were calculated using a formula specific to Daymet estimates containing T_max_, Tmin, VP, and three constants, as described in prior literature [14].

Death certificates for Chicago decedents are maintained by the Illinois Department of Public Health and provided by the Chicago Department of Public Health. In addition to decedent demographic characteristics like sex, race, date of death, and date of birth, we retrieved the geospatial markers of decedent residential address and community area. Each decedent had one primary and up to three contributing causes of death with their respective International Classification of Diseases (ICD-10) codes listed on their death certificate. A CVD cause of death was classified as such if any primary or contributory causes of death contained an ICD-10 code between I00 and I99. The following CVD subtypes were classified via ICD-10 codes as: coronary heart disease (CHD) I20-I25, stroke I60-I69, and myocardial infarction (MI) I21-I22, per American Heart Association guidelines [15].

The Chicago Area Patient-Centered Outcomes Research Network (CAPriCORN) is a collaborative network of healthcare institutions and community organizations that leverages electronic health record data from participating sites as part of the larger Patient-Centered Outcomes Research Network framework. CAPriCORN has a patient population of nearly 13 million and has been extensively used in prior health outcomes research [16, 17]. We queried all ED encounters across all participating CAPriCORN institutions from 2011-2022, which included encounter-specific ICD-10 diagnosis codes, patient demographic information, and patient residential address census tract. We joined each ED encounter to its respective community area using a spatial join of the patient’s residential address census tract. CVD- and CVD subtype-related ED encounters were identified using the same ICD-10 codes as listed above for CVD mortality.

### Statistical Analysis

We selected four different temporal scales for this study: annual, monthly, daily, and daily lag (3-day rolling average). We aggregated CVD mortalities and ED encounters by community area and calculated the respective rate per 100,000 population by temporal scale (annual, monthly, daily, and daily lag). Mean T_max_ and RH were also aggregated by community area and temporal scale. Annual ACS estimates were joined by year and community area with the assumption that these estimates remained the same for each respective year.

#### Generalized Additive Model Heat-Related Excess Morbidity and Mortality Estimates

The generalized additive model (GAM) is a robust framework for regressing non-linear predictors against health outcomes and numerous studies have utilized GAMs for environmental health impact research [18–21]. It is well-established that the relationship between temperature and CVD outcomes is nonlinear with a “U” shaped curve, which can be modeled in a variety of ways [22, 23]. GAMs can accommodate different temporal scales and lag structures, handle nonlinear multidimensional interactions with tensor smooths, and have improved interpretability over methods like distributed lag nonlinear models [24, 25].

We selected final GAM covariates based on Akaike Information Criterion and Bayesian Information Criterion values. Final covariates included RH, median age, median income, education level, racial composition, employment status, sex distribution, and total population (offset). The primary exposure was T_max_. Overdispersion was measured for each model using the deviance divided by the residual degrees of freedom with a value ≥1 indicating overdispersion. We employed 10-fold cross-validation for four GAMs addressing different cumulative heat exposure temporal scales: annual, monthly, daily, and daily lag. All GAMs had a negative binomial link function to address overdispersion in the CVD outcome rates, while smooth functions were incorporated to model nonlinear associations for temperature, RH, and specific sociodemographic covariates. To account for seasonality and long-term temporal trends, cubic regression spline functions for the year (annual models), month (monthly models), or date (daily and daily lag models) were included in each model.

The GAMs provided estimated CVD mortality or ED visit (morbidity) rates per 100,000 population for annual, monthly, daily, and daily lag temporal scales by community area. Using the entire range of temperatures within each respective temporal scale, we identified the “peak threshold” (the temperature at which heat-related excess CVD mortality or ED visit rates are maximized across all 77 community areas) and “highest positive threshold” (the highest temperature threshold that still results in positive heat-related excess CVD mortality or ED visit rates across all 77 community areas). To calculate heat-related excess CVD mortality or ED visit rates by community area, the cumulative below-threshold mean estimated CVD mortality or ED visit rate was subtracted from the cumulative above-threshold mean estimated CVD mortality or ED visit rate. To quantify uncertainty, we used bootstrapping (1,000 samples) to estimate 95% confidence intervals for HRCVD mortality and ED visit rates at each temperature threshold.

#### Principal Component Analysis Clustering of Chicago Community Areas

For models with statistically significant heat-related excess rates, we investigated spatiotemporal patterning by community area. Using the respective calculated peak temperature thresholds for annual, monthly, and daily temporal scales, we standardized heat-related excess mortality rates per 100,000 population using z-score normalization to ensure community area comparability across temporal scales.

We conducted principal component analysis (PCA) on standardized heat-related excess mortality rates across three temporal scales: annual, monthly, and daily. This dimensionality reduction approach transformed these correlated metrics into orthogonal principal components (PCs), each representing a distinct pattern of variability in heat-related mortality across communities. We interpreted the resulting PC loadings to characterize the contribution of each original variable to each principal component. PC1 and PC2 were retained based on the proportion of variance explained and loading strength, with PC1 typically reflecting either long-term (yearly/monthly) or acute (daily) excess mortality patterns, depending on the direction and magnitude of loadings.

Each community was assigned a score along PC1 and PC2, representing its position in the transformed mortality space. We then applied k-means clustering to the PC scores to group communities into phenotypic categories based on shared temporal mortality patterns. Clusters were labeled based on their centroid positions in PC space and the dominant mortality profiles they represented. Final cluster definitions and naming conventions were based on the directionality of loadings and spatial separation in the biplot, and are further detailed in Supplemental Files.

We applied K-means clustering on community areas using the Elbow Method to determine the optimal number of clusters. Cluster assignments were then overlaid onto a PCA biplot to visualize how community areas group along these two principal dimensions for heat-related excess rate vulnerability indexing.

### Sensitivity Analysis

We performed a sensitivity analysis to assess the agreement of peak and highest positive temperature thresholds obtained from estimating heat-related excess mortality and ED visit rates with the GAMs, with the corresponding thresholds calculated using crude observed heat-related excess mortality and ED visit rates. These comparisons were conducted for CVD and its subtypes across all 77 community areas at each temporal scale.

We performed all geospatial and statistical analyses using R (R Core Development Team 2024, v4.3.2) [26]. Geoanalytic and mapping packages included ‘daymetr’, ‘exactextractr’, ‘sf’, ‘terra’, and ‘tmap’ [27–31]. The package ‘tidycensus’ queried ACS tables and variables, ‘mgcv’ provided the GAM functions, while ‘dplyr’, ‘ggplot2’, ‘lubridate’, ‘zoo’, ‘boot’, ‘cluster’, ‘factoextra’, and ‘caret’ were used for data processing, analysis, and figure creation [32–40].

## RESULTS

### Study Cohort

From May to September 2011-2022, there were an initial 96,791 adult decedents and 782,416 ED visits within 325,530 unique adult patients with a Chicago address at the time of event. This yielded a total of 46,843 (48.4%) decedents with a CVD cause of death (50.9% male, 49.1% female; mean [standard deviation] age: 73.7 years [15.6 years]) and 219,044 (28.0%) total ED visits with a CVD diagnosis (44.5% male, 55.5% female; mean [standard deviation] age: 59.4 years [16.9 years]) during the study period, all of which were included in the rest of the analysis. Among CVD deaths, 33.3% were CHD deaths, followed by stroke (15.9%), and MI (10.7%). Among CVD ED visits, 17.1% were CHD ED visits, followed by stroke (8.9%), and MI (2.1%). The mean (standard deviation) May-September temperature was 26.07°C (0.73°C) and the mean (standard deviation) relative humidity was 67.7% (1.60%) on the day of death or ED visit. Table 1 provides the demographic characteristics of the CVD cohorts and Figure 1 displays the spatial distributions of temperature, RH, CVD deaths, and CVD ED visits.

**Figure 1:**
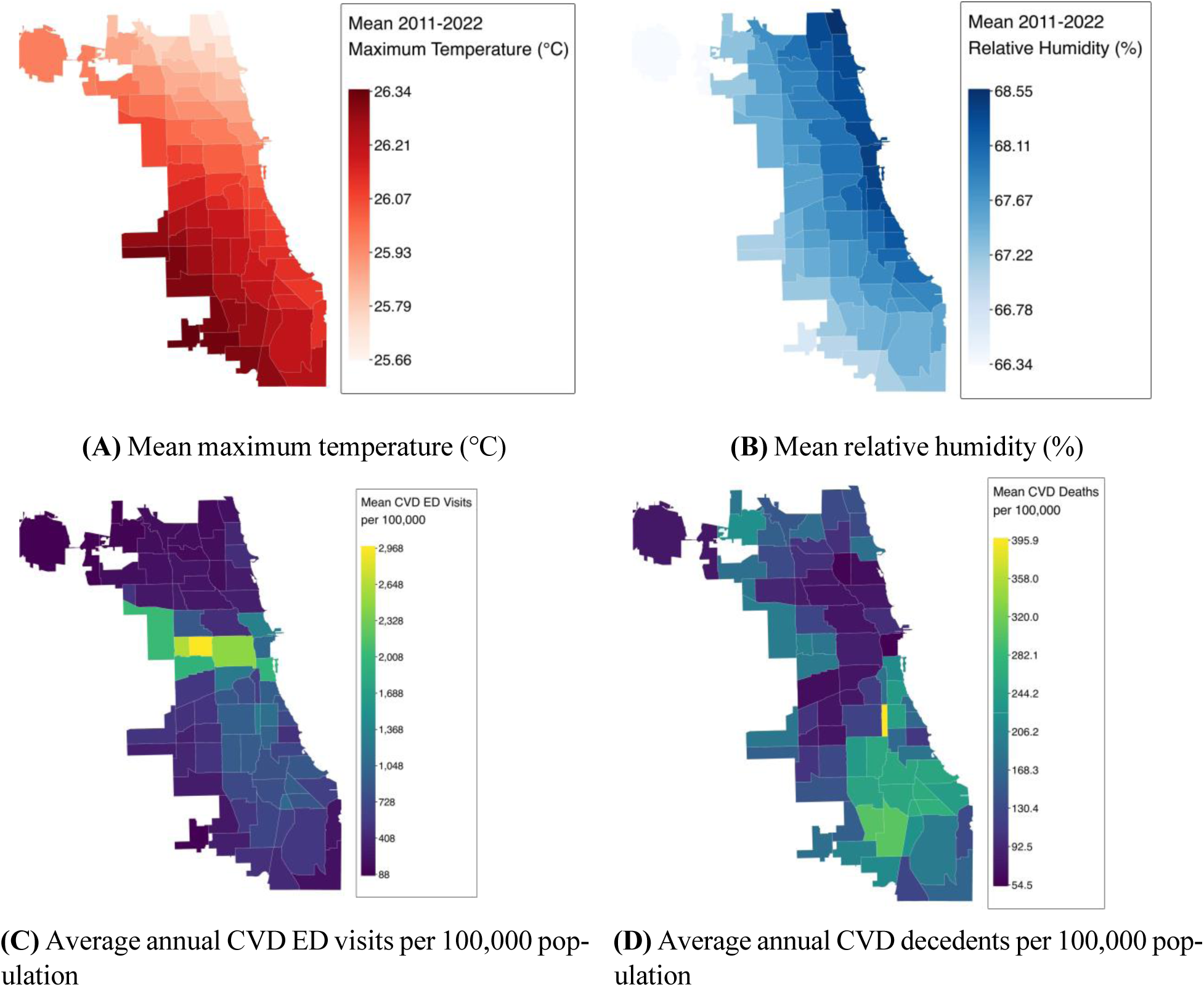
Mean maximum temperature (top left), relative humidity (top right), and annual CVD morbidity (bottom left) and mortality (bottom right) rates per 100,000 population by Chicago community area from May to September (2011 to 2022). CVD = cardiovascular disease; ED = emergency department.

**Table 1:** Total number of adult CVD deaths and ED visits across all Chicago community areas during warm months (May-September) from 2011-2022. CVD = cardiovascular disease; ED = emergency department; IQR = interquartile range.

| Category | CVD Deaths<br>(n = 46,843) | CVD ED Visits<br>(n = 219,044) | Median (IQR)<br>CVD Deaths<br>across all<br>community<br>areas | Median (IQR)<br>CVD ED Visits<br>across all<br>community<br>areas |
| --- | --- | --- | --- | --- |
| **Race** |  |  |  |  |
| Asian | 1376 (2.9%) | 3370 (1.5%) | 7 (2–25) | 19 (5–58) |
| Black Hispanic | 318 (0.7%) | 792 (0.4%) | 4 (2–7.25) | 8.5 (3–16) |
| Black non-Hispanic | 22,985 (49.1%) | 105,073 (48%) | 126.5<br>(26.25–429) | 566 (213–1317) |
| Latino/a | 879 (1.9%) | 19,833 (9.1%) | 7 (2–20) | 124 (52–301) |
| Other | 328 (0.7%) | 59,341 (27.1%) | 3 (1–7) | 515 (243–806) |
| White Hispanic | 4890 (10.4%) | 13,066 (6%) | 34.5 (5–92.25) | 72 (24–202) |
| White non-Hispanic | 16,067 (34.3%) | 17,569 (8%) | 124.5<br>(18.75–301) | 143 (62–316) |
| **Age** |  |  |  |  |
| 65 years and above | 34,148 (72.9%) | 83,582 (38.2%) | 365 (250–573) | 675 (367–1177) |
| Below 65 years | 12,682 (27.1%) | 135,462 (61.8%) | 124 (72–220) | 1185<br>(556–1974) |
| **Sex** |  |  |  |  |
| Female | 22,988 (49.1%) | 121,639 (55.5%) | 249 (158–400) | 967 (514–1816) |
| Male | 23,855 (50.9%) | 97,375 (44.5%) | 282 (161–432) | 852 (448–1520) |

### Generalized Additive Model Results

We applied cubic regression splines as GAM smoothing functions to T_max_, median age, median income, and relative humidity, as well as to the time variable (year, month, or date) to account for seasonality and long-term trends. All other sociodemographic covariates were modeled as linear terms. The sociodemographic covariates at the community level which were included in the GAMs are displayed by study period mean T_max_ tertile in Table 2.

**Table 2:** American Community Survey (ACS) estimates across Chicago community areas stratified by maximum temperature (T_max_) tertiles over the study period; Low (25.66°C to 25.99°C), Medium (26.01°C to 26.16°C), and High (26.17°C to 26.34°C). IQR = interquartile range.

| ACS Variable | Category | Low $T_{\max}$ Tertile | Medium $T_{\max}$ Tertile | High $T_{\max}$ Tertile |
| --- | --- | --- | --- | --- |
|  | Total Population | 53,206.1<br>(32,122.7–66,681.4) | 21,032.1<br>(14,210.2–32,778.4) | 28,255.1<br>(18,896.9–38,017) |
|  | Median Age (Years) | 34.9 (32.5–38.1) | 34.3 (31.6–37.3) | 36.3 (32.3–39.7) |
| | Median Income (\$) | 64,687.4<br>(54,541.3–92,740.4) | 33,661.5<br>(28,336.2–50,022.8) | 41,928.6<br>(33,791.5–53,292.8) |
| Education (%) | College | 19.2 (14.9–27.5) | 9.4 (6.1–13.3) | 5.9 (4.3–9.1) |
|  | High School | 12.2 (8.1–15.8) | 15.1 (11.9–18.3) | 18.1 (15.7–20) |
| Race (%) | White | 73.1 (62.4–80.2) | 11.1 (4.2–44.5) | 30.2 (2.8–57.2) |
|  | Black | 3.7 (2.2–10.2) | 76 (23.2–92.2) | 50.6 (5.5–94.6) |
|  | Latino | 19 (13.3–42.2) | 4.9 (3–21.8) | 9.3 (3.3–53.8) |
|  | Asian | 6.6 (4.5–11.2) | 0.6 (0.3–10.5) | 0.5 (0.3–1) |
| Employment (%) | Employed | 53.7 (49.8–61.8) | 38.9 (35–46.4) | 39.7 (35.7–43.8) |
|  | Unemployed | 4 (3.4–4.5) | 7.2 (5.9–8.9) | 8.2 (5.6–9.6) |
| Sex (%) | Male | 49.2 (48.3–50.4) | 45.6 (43.5–47.8) | 48 (45.4–49.5) |
|  | Female | 50.1 (49.2–50.9) | 53.6 (51.8–55.4) | 52 (50.1–54.3) |

Maximum temperature was significantly associated with heat-related excess CVD mortality rate for all temporal scales (annual, monthly, daily, and daily lag (0-3 days)), and significantly associated with heat-related excess CHD mortality rate for annual, monthly, and daily, but not daily lag (0-3 days), temporal scales. Heat-related excess MI and stroke mortality rates had no significant association with T_max_ except for MI during the daily lag (0-3 days) temporal scale, where temperature had a statistically significant protective effect. Though trends were observed, there were no statistically significant relationships between T_max_ and heat-related excess ED visit rates for any temporal scale or CVD subtype, apart from annual heat-related excess MI ED visit rates, which likely is spurious. Using the GAM estimates, peak and highest positive temperature thresholds were identified and reported in Table 3 (mortality) and Table 3.4-3.5 (ED visits). Although no statistically significant relationship emerged for cardiovascular ED visits and temperature at any temporal scale, inverted spatial patterns were observed when compared to cardiovascular mortality, which suggests that populations with higher heat-related excess cardiovascular ED visit rates may not overlap with populations with greater heat-related cardiovascular mortality vulnerability (Supplemental Figure 1).

**Table 3:** Average heat-related excess cardiovascular mortality rate across all 77 Chicago community areas at peak and highest positive thresholds from May to September (2011 to 2022). Values with “NA” indicate that there were no thresholds where the average heat-related excess mortality rate exceeded 0. CVD = cardiovascular disease; CI = confidence interval; T_max_ = maximum temperature; CHD = coronary heart disease; MI = myocardial infarction.

| Mortality | Temporal Scale | Peak Threshold (°C \|\| °F) | Peak Avg Excess Rate per 100,000 (95% CI) | Highest Positive Threshold (°C \|\| °F) | Highest Positive Avg Excess Rate per 100,000 (95% CI) | P-value |
| --- | --- | --- | --- | --- | --- | --- |
| CVD | Annual | 25.9°C \|\| 78.6°F | 23.00 (19.80, 26.50) | 26.4°C \|\| 79.5°F | 2.63 (-2.28, 6.97) | <0.001 |
|  | Monthly | 29.4°C \|\| 84.9°F | 2.02 (1.38, 2.62) | 30.2°C \|\| 86.4°F | 1.09 (0.23, 1.96) | <0.001 |
|  | Daily | 36.0°C \|\| 96.8°F | 0.026 (0.016, 0.037) | 39.0°C \|\| 102.2°F | 0.016 (-0.009, 0.041) | <0.001 |
|  | Daily lag (0–3) | 38.0°C \|\| 100.4°F | 0.109 (0.084, 0.139) | 38.5°C \|\| 101.3°F | 0.108 (0.081, 0.144) | <0.001 |
| CHD | Annual | 27.4°C \|\| 81.3°F | 9.06 (7.24, 10.60) | 27.7°C \|\| 81.9°F | 7.80 (4.12, 12.10) | 0.031 |
|  | Monthly | 31.7°C \|\| 89.1°F | 3.30 (2.91, 3.66) | 32.7°C \|\| 90.9°F | 3.08 (2.54, 3.74) | 0.0013 |
|  | Daily | 38.5°C \|\| 101.3°F | 0.098 (0.092, 0.104) | 39.0°C \|\| 102.2°F | 0.093 (0.081, 0.105) | 0.0374 |
|  | Daily lag (0–3) | 38.0°C \|\| 100.4°F | 0.121 (0.111, 0.131) | 38.5°C \|\| 101.3°F | 0.119 (0.105, 0.133) | 0.16 |
| MI | Annual | 25.5°C \|\| 77.9°F | 0.56 (0.13, 0.87) | 26.5°C \|\| 79.7°F | 0.03 (-0.42, 0.38) | 0.854 |
|  | Monthly | 32.4°C \|\| 90.3°F | 0.33 (0.21, 0.43) | 32.7°C \|\| 90.9°F | 0.27 (0.10, 0.49) | 0.201 |
|  | Daily | 31.5°C \|\| 88.7°F | -0.003 (-0.003, -0.003) | NA \|\| NA | NA | 0.185 |
|  | Daily lag (0–3) | 30.5°C \|\| 86.9°F | -0.006 (-0.006, -0.006) | NA \|\| NA | NA | 0.0422 |
| Stroke | Annual | 25.6°C \|\| 78.1°F | 4.50 (3.97, 4.88) | 26.8°C \|\| 80.2°F | 0.13 (-0.76, 0.83) | 0.1491 |
|  | Monthly | 28.0°C \|\| 82.4°F | 0.01 (-0.04, 0.07) | 28.0°C \|\| 82.4°F | 0.01 (-0.04, 0.07) | 0.2369 |
|  | Daily | 24.5°C \|\| 76.1°F | -0.004 (-0.004, -0.003) | NA \|\| NA | NA | 0.2464 |
|  | Daily lag (0–3) | 25.0°C \|\| 77.0°F | -0.004 (-0.004, -0.004) | NA \|\| NA | NA | 0.6097 |

**Table 4:** Average heat-related excess cardiovascular emergency department (ED) visit rate across all 77 Chicago community areas at peak and highest positive thresholds from May to September (2011 to 2022). Values with “NA” indicate that there were no thresholds where the average heat-related excess ED visit rate exceeded 0. CVD = cardiovascular disease; CI = confidence interval; T_max_ = maximum temperature; CHD = coronary heart disease; MI = myocardial infarction.

| ED Visits | Temporal Scale | Peak Threshold (°C \|\| °F) | Peak Avg Excess Rate per 100,000 (95% CI) | Highest Positive Threshold (°C \|\| °F) | Highest Positive Avg Excess Rate per 100,000 (95% CI) | P-value |
| --- | --- | --- | --- | --- | --- | --- |
| CVD | Annual | 25.6°C \|\| 78.1°F | -8.95 (-40.80, 21.30) | NA \|\| NA | NA | 0.8213 |
|  | Monthly | 20.2°C \|\| 68.4°F | 12.40 (7.51, 18.30) | 26.4°C \|\| 79.5°F | 0.81 (-1.58, 3.07) | 0.585 |
|  | Daily | 20.5°C \|\| 68.9°F | 0.134 (0.115, 0.155) | 26.0°C \|\| 78.8°F | 0.022 (0.008, 0.036) | 0.0842 |
|  | Daily lag (0–3) | 21.0°C \|\| 69.8°F | 0.176 (0.156, 0.195) | 25.5°C \|\| 77.9°F | 0.010 (-0.004, 0.025) | 0.181 |
| CHD | Annual | 25.7°C \|\| 78.3°F | -3.20 (-8.66, 1.69) | NA \|\| NA | NA | 0.1754 |
|  | Monthly | 20.2°C \|\| 68.4°F | 2.50 (1.82, 3.20) | 27.0°C \|\| 80.6°F | 0.03 (-0.51, 0.46) | 0.487 |
|  | Daily | 20.5°C \|\| 68.9°F | 0.028 (0.025, 0.032) | 26.5°C \|\| 79.7°F | 0.005 (0.002, 0.006) | 0.219 |
|  | Daily lag (0–3) | 21.0°C \|\| 69.8°F | 0.036 (0.032, 0.039) | 26.0°C \|\| 78.8°F | 0.002 (-0.001, 0.006) | 0.356 |
| MI | Annual | 25.5°C \|\| 77.9°F | 3.65 (2.90, 4.55) | 26.2°C \|\| 79.2°F | 0.80 (-0.29, 1.92) | <0.001 |
|  | Monthly | 26.4°C \|\| 79.5°F | 0.30 (0.22, 0.38) | 29.4°C \|\| 84.9°F | 0.153 (-0.03, 0.33) | 0.83 |
|  | Daily | 23.0°C \|\| 73.4°F | 0.004 (0.004, 0.005) | 29.5°C \|\| 85.1°F | 0.001 (0.000, 0.001) | 0.734 |
|  | Daily lag (0–3) | 21.5°C \|\| 70.7°F | 0.006 (0.005, 0.006) | 29.0°C \|\| 84.2°F | 0.001 (-0.001, 0.001) | 0.494 |
| Stroke | Annual | 26.0°C \|\| 78.8°F | 9.33 (6.15, 13.20) | 26.2°C \|\| 79.2°F | 0.134 (-3.98, 3.91) | 0.702 |
|  | Monthly | 20.2°C \|\| 68.4°F | 1.70 (1.19, 2.35) | 27.7°C \|\| 81.9°F | 0.01 (-0.23, 0.41) | 0.579 |
|  | Daily | 22.5°C \|\| 72.5°F | 0.019 (0.017, 0.022) | 28.0°C \|\| 82.4°F | 0.003 (0.001, 0.005) | 0.262 |
|  | Daily lag (0–3) | 21.5°C \|\| 70.7°F | 0.029 (0.027, 0.031) | 27.5°C \|\| 81.5°F | 0.002 (0.001, 0.003) | 0.188 |

#### Cardiovascular Disease Mortality

When the mean annual T_max_ (May-September) reaches 78.6°F, there were an estimated 23 heat-related excess CVD mortalities per 100,000 population per year (95% CI: 19.80-26.50, p*<*0.001). When the mean monthly T_max_ reaches 84.9°F, there were an estimated 2.02 heat-related excess CVD mortalities per 100,000 population per month (95% CI: 1.380-2.620, p*<*0.001). When daily T_max_ reaches 96.8°F, there were an estimated 0.0262 heat-related excess CVD mortalities per 100,000 population per day (95% CI: 0.0164-0.0370, p*<*0.001). A 3-day average T_max_ of 100.4°F corresponded to a heat-related excess rate of 0.1090 CVD mortalities per 100,000 population per day (95% CI: 0.0844-0.1390, p*<*0.001). Temperature curves and spatial distributions can be found in Figures 2 to 4. Annual mean temperature had a positive linear relationship to annual heat-related excess CVD mortality rates, while monthly and daily temperatures had a U-shaped non-linear relationship to their respective excess mortality rates. All smoothed GAM covariates can be found in Supplemental Files.

**Figure 2:**
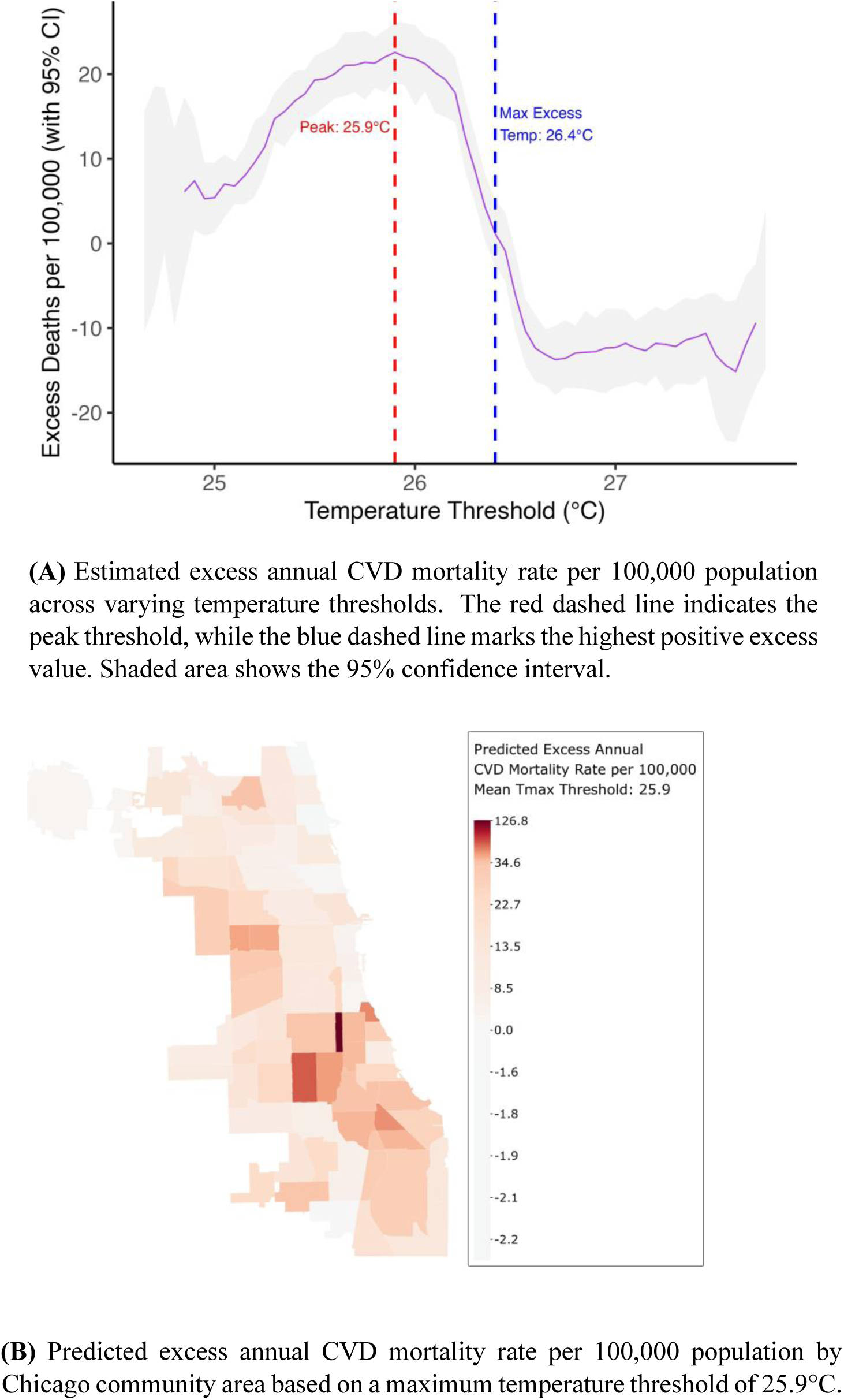
Temperature-based excess CVD mortality estimates: (A) modeled excess annual mortality across T_max_ thresholds, and (B) spatial distribution of excess annual CVD deaths across Chicago community areas.

**Figure 3:**
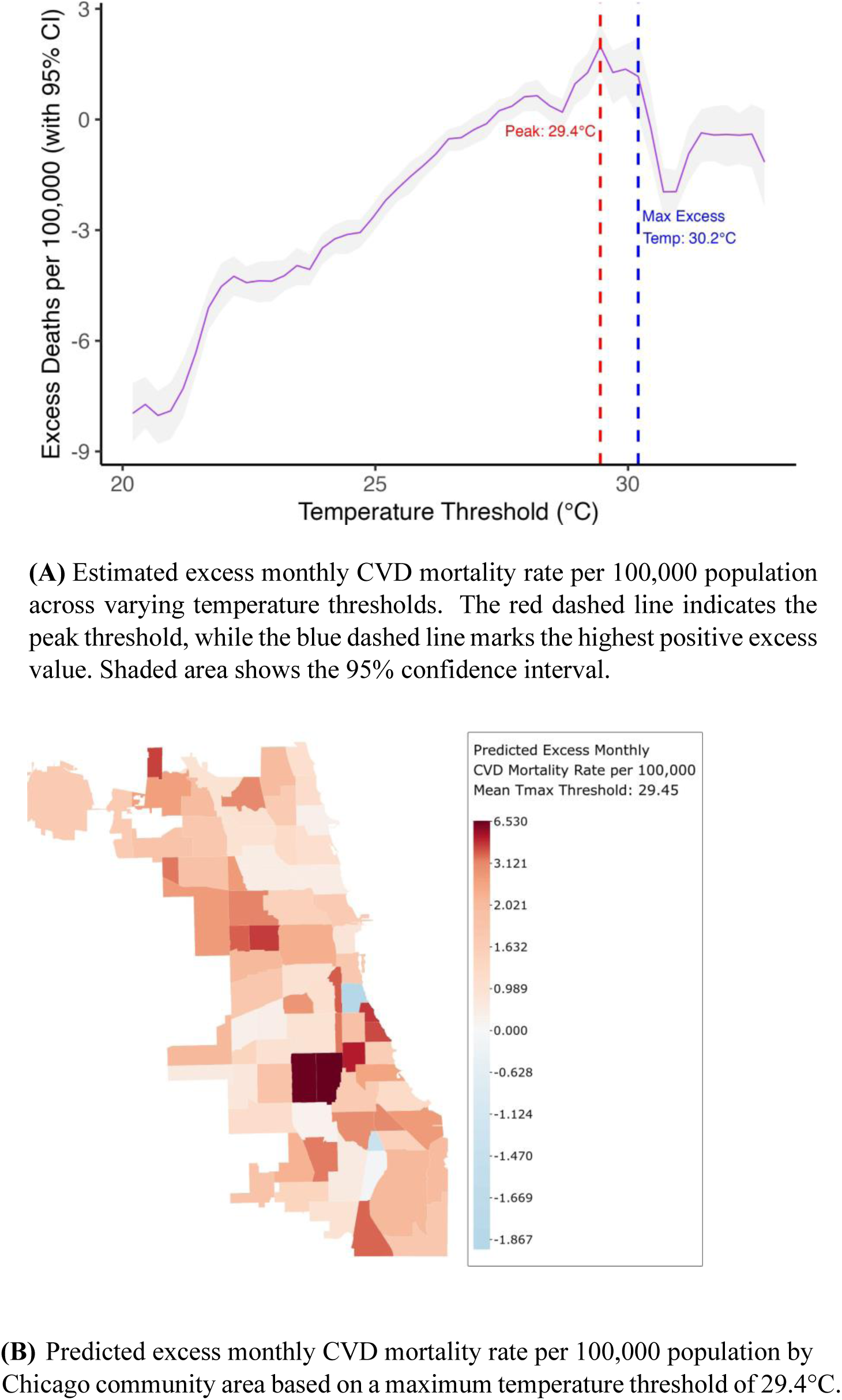
Temperature-based excess CVD mortality estimates: (A) modeled excess monthly mortality across T_max_ thresholds, and (B) spatial distribution of excess monthly CVD deaths across Chicago community areas.

**Figure 4:**
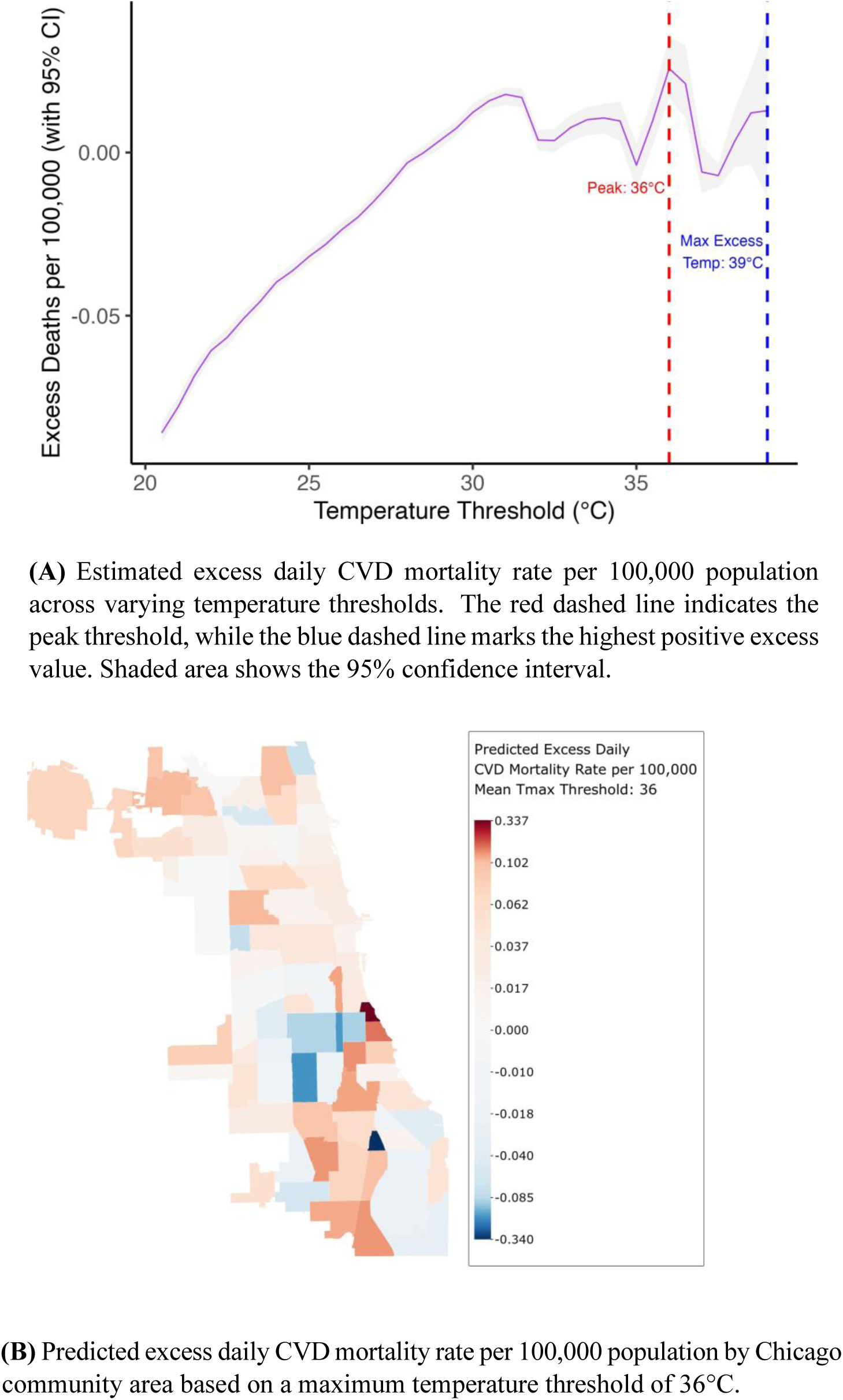
Temperature-based excess CVD mortality estimates: (A) modeled excess daily mortality across T_max_ thresholds, and (B) spatial distribution of excess daily CVD deaths across Chicago community areas.

Higher T_max_ consistently predicted increased heat-related excess CVD mortality across all four temporal scales, with the strongest associations observed at the monthly (p*<*0.001) and daily levels (p*<*0.001), with the 0–3-day lag period further strengthening the daily association. Humidity was a significant predictor annually (p=0.003), but its explanatory power decreased at monthly (p=0.35) and daily (p=0.12) temporal scales. Increased median age and decreased income remained significant predictors (p*<*0.001) for increased heat-related excess CVD mortality at all temporal scales. Community areas with higher proportions of Black residents were linked to increased heat-related excess CVD mortality for monthly (p=0.04) and daily (p=0.02) temporal scales. A higher proportion of Hispanic residents was associated with lower heat-related excess CVD mortality (p*<*0.001), as was a higher proportion of male residents (p=0.04). The significant predictors of the CVD mortality models and their estimated effects can be found in Supplemental Files.

#### Coronary Heart Disease Mortality

When the mean annual T_max_ (May-September) reaches 81.3°F, there were an estimated 9.06 heat-related excess CHD mortalities per 100,000 population per year (95% CI: 7.24-10.60, p=0.031). When the mean monthly T_max_ reaches 89.1°F, there were an estimated 3.30 heat-related excess CHD mortalities per 100,000 population per month (95% CI: 2.91-3.66, p=0.0013). When daily T_max_ reaches 101.3°F, there were an estimated 0.0979 heat-related excess CHD mortalities per 100,000 population per day (95% CI: 0.0919-0.1040, p=0.037). A 3-day average T_max_ of 100.4°F corresponded to a heat-related excess rate of 0.1210 CHD mortalities per 100,000 population per day (95% CI: 0.1110-0.1310, p=0.16). Temperature curves and spatial distributions can be found in Figures 5 to 7. Annual mean temperature had a positive linear relationship to annual excess CHD mortality rate, while monthly and daily temperatures had a U-shaped non-linear relationship to their respective heat-realted excess CHD mortality rates. All smoothed GAM covariates can be found in Supplemental Files.

**Figure 5:**
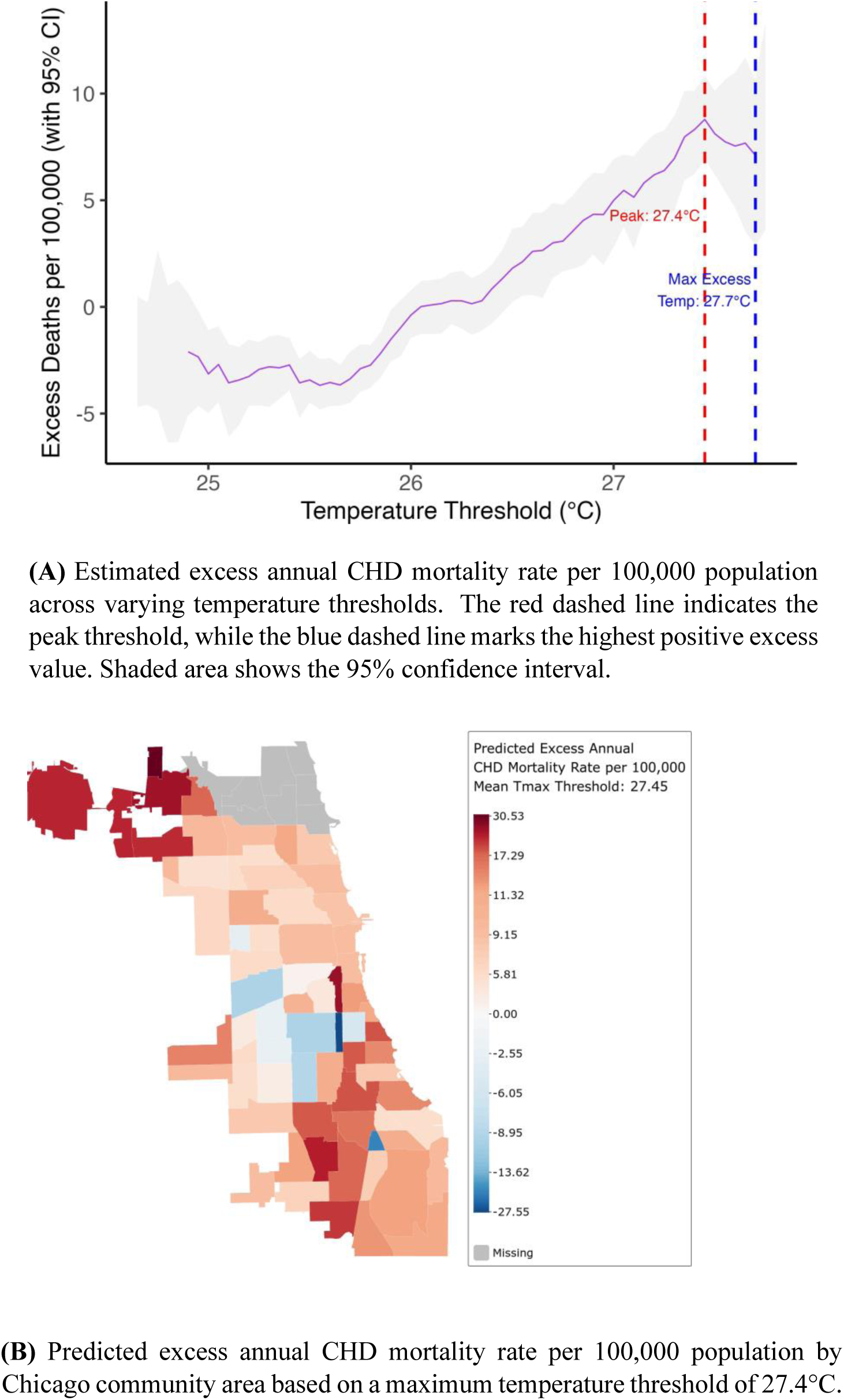
Temperature-based excess CHD mortality estimates: (A) modeled excess annual mortality across T_max_ thresholds, and (B) spatial distribution of excess annual CHD deaths across Chicago community areas.

**Figure 6:**
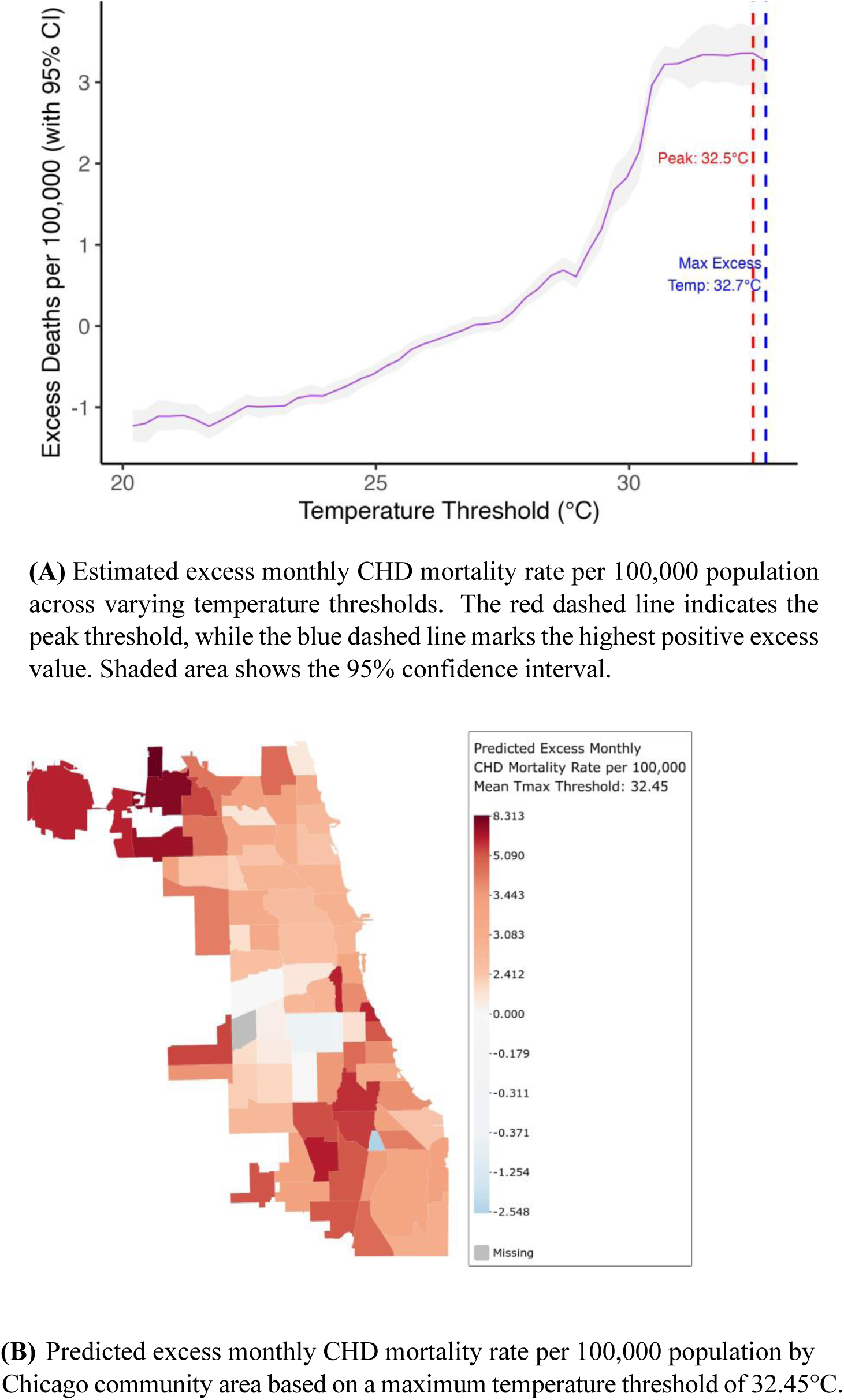
Temperature-based excess CHD mortality estimates: (A) modeled excess monthly mortality across T_max_ thresholds, and (B) spatial distribution of excess monthly CVD deaths across Chicago community areas.

**Figure 7:**
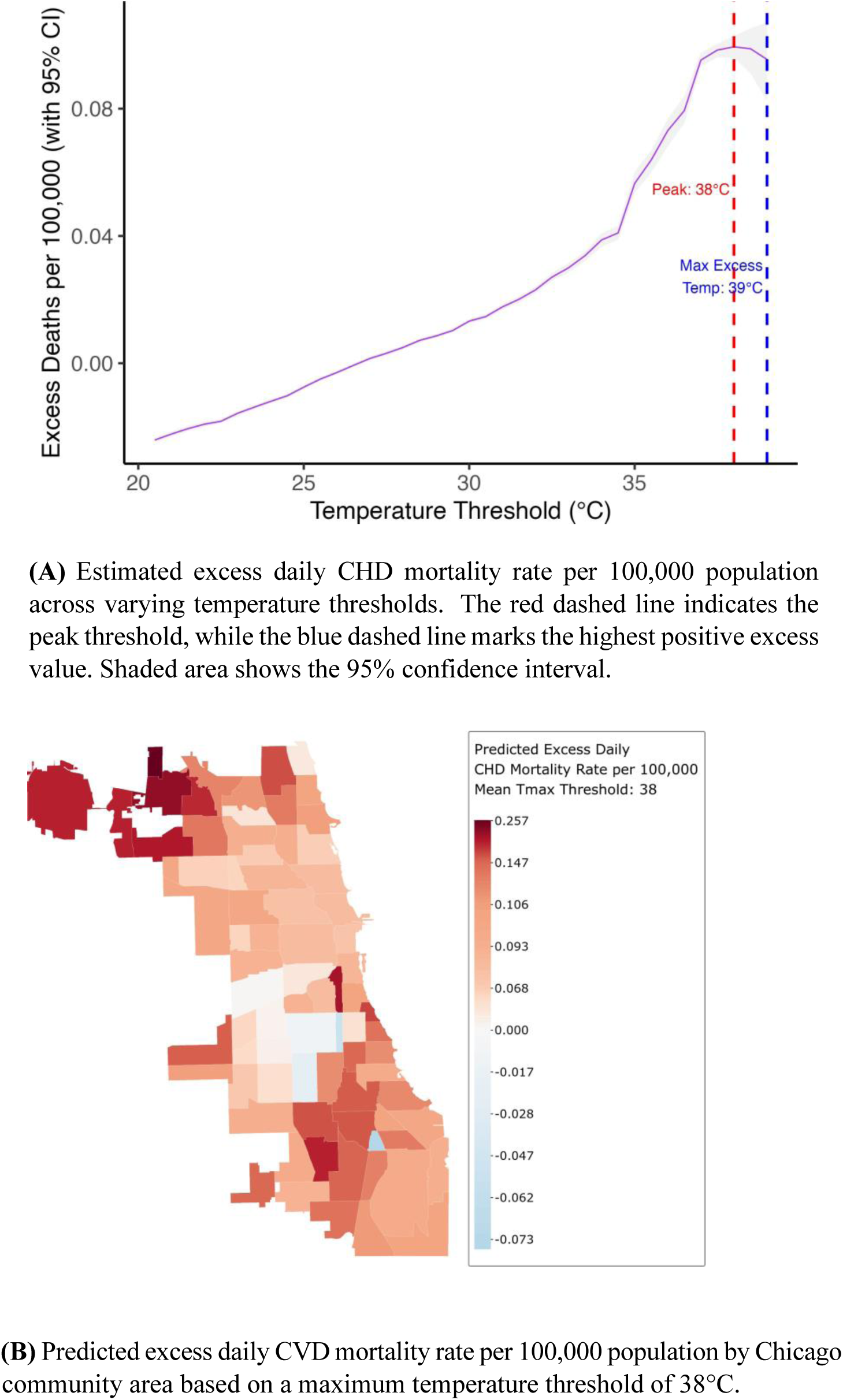
Temperature-based excess CHD mortality estimates: (A) modeled excess daily mortality across T_max_ thresholds, and (B) spatial distribution of excess daily CHD deaths across Chicago community areas.

Patterns for heat-related excess CHD mortality were similar to CVD. Higher T_max_ was significantly predictive of increased heat-related excess CHD mortality for all temporal scales except daily lag 0-3 (p=0.16) and RH was only significant for the annual temporal scale (p=0.02). Predictors such as lower median income, higher median age, and higher percentage of residents with a high school education were associated with increased heat-related excess CHD mortality for all temporal scales. A higher proportion of Hispanic residents was associated with lower heat-related excess CHD mortality, as was a higher proportion of Asian residents at all temporal scales (p*<*0.001). The significant predictors of the CHD mortality models and their estimated effects can be found in Supplemental Files.

### Principal Component Analysis Clustering for Long-Term Versus Short-Term Vulnerability

To better understand spatial patterns of heat-related excess CVD and CHD mortality rates in Chicago, we conducted a PCA and applied K-means clustering to classify community areas based on their vulnerability to short-term (daily) and long-term (monthly/annual) heat-related excess cardiovascular mortality during extreme heat. Figures 8 and 9 display the results of this analysis, with PCA biplots illustrating the clustering structure (A, top panels) and corresponding spatial distributions of vulnerability categories across Chicago community areas (B, bottom panels).

**Figure 8:**
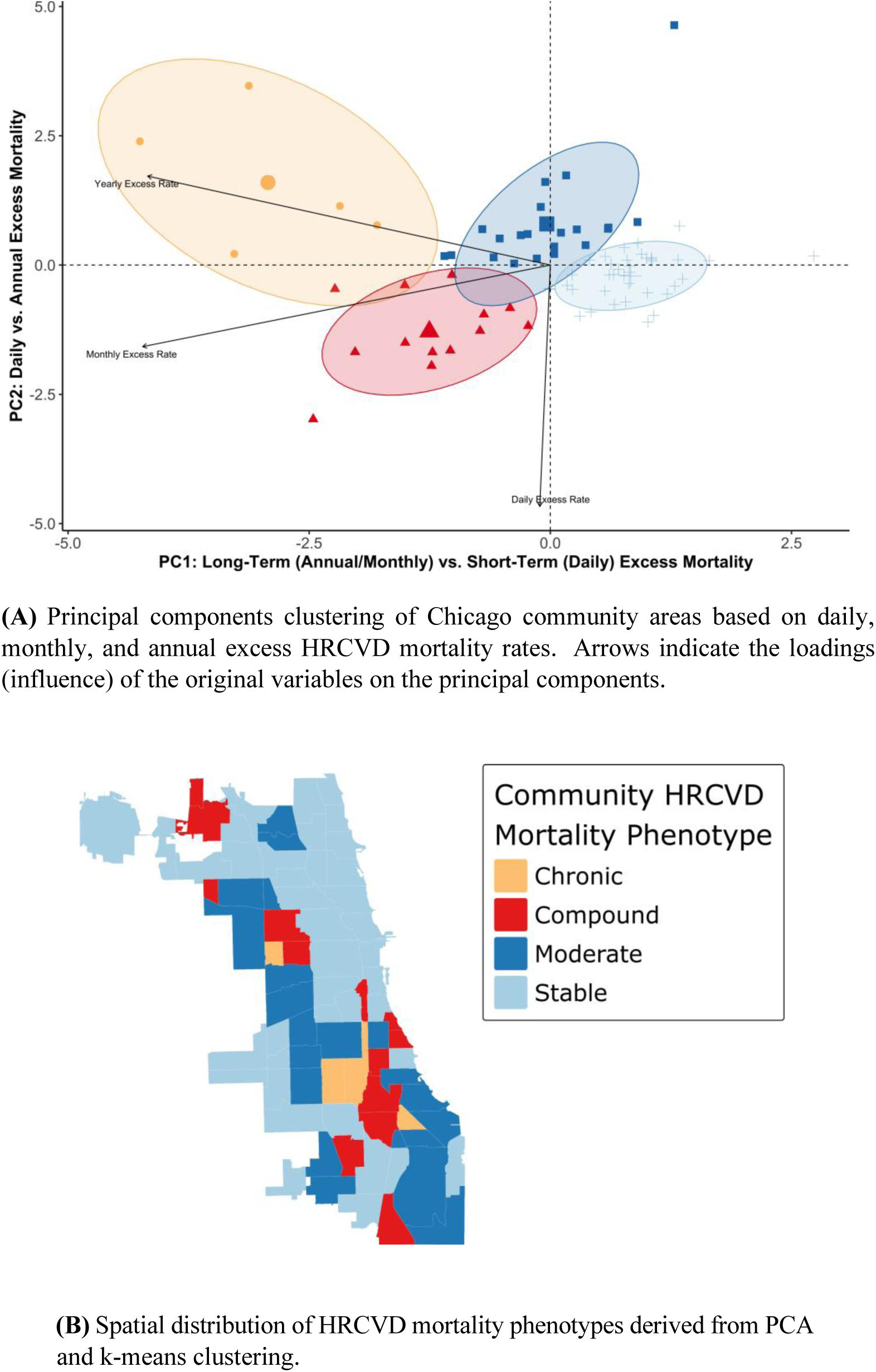
PCA-derived CVD vulnerability clusters based on temporal patterns of excess mortality (daily, monthly, and annual) and their spatial distribution across Chicago community areas from May to September (2011–2022). PCA = principal component analysis; HRCVD = heat-related cardiovascular disease.

**Figure 9:**
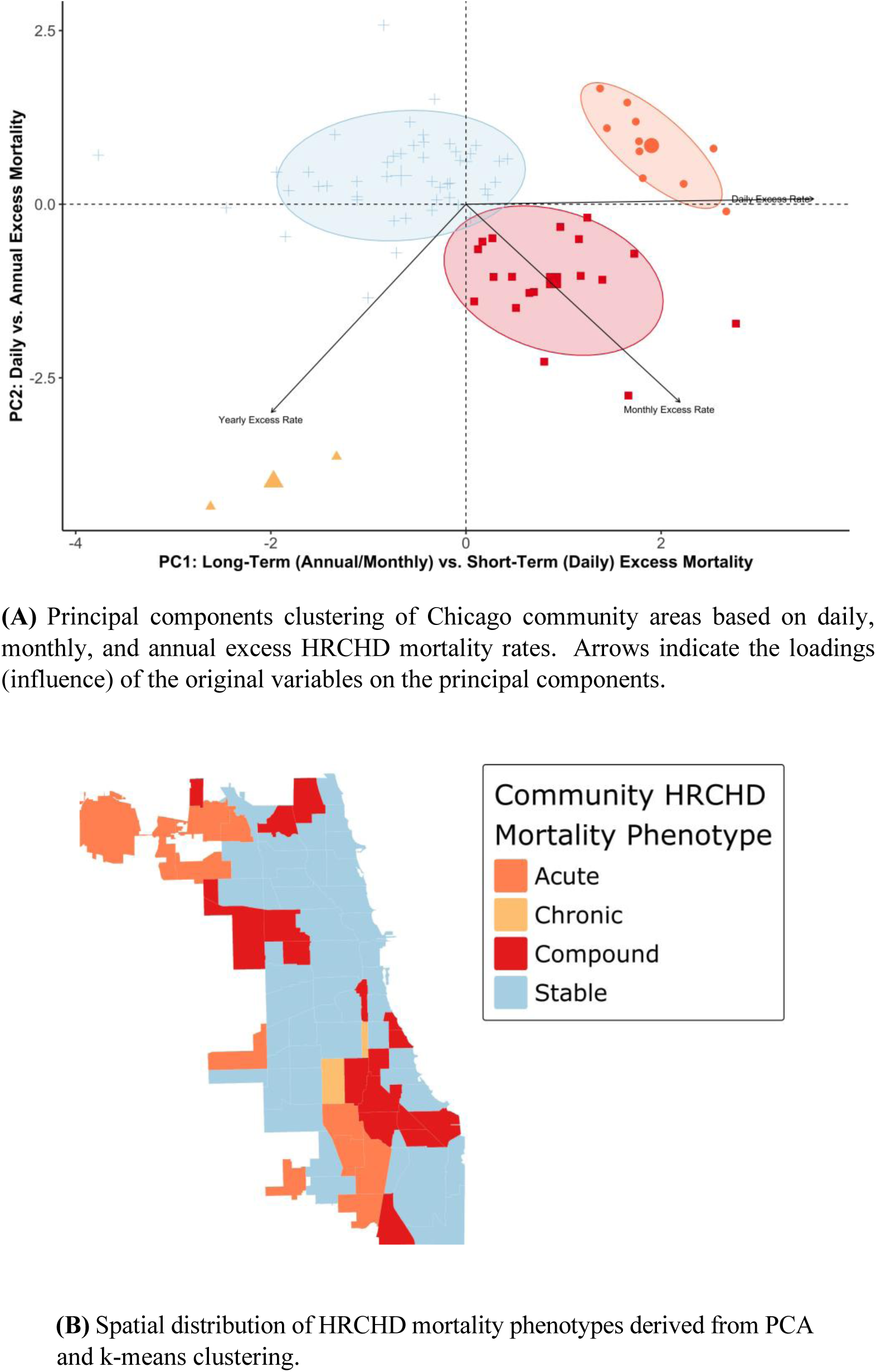
PCA-derived HRCHD vulnerability clusters based on temporal patterns of excess mortality (daily, monthly, and annual) and their spatial distribution across Chicago community areas from May to September (2011–2022). PCA = principal component analysis; HRCHD = heat-related coronary heart disease.

Our analysis identified five distinct clusters of community-level vulnerability to HRCVD and HRCHD based on principal component scores derived from annual, monthly, and daily excess mortality rates. Stable communities exhibited low excess mortality across all temporal scales, indicating minimal vulnerability to both acute and long-term heat exposure. Acute Vulnerability communities experienced elevated mortality primarily in response to short-term (daily) temperature extremes, with limited evidence of chronic exposure effects. Chronic Vulnerability communities showed sustained excess mortality associated with longer-term (monthly and annual) heat patterns, but minimal daily spikes. Moderate Vulnerability communities exhibited modest excess mortality levels, particularly from chronic exposure, without clear dominance of acute or long-term risk. Compound Vulnerability communities demonstrated consistently high heat-related excess mortality across both acute and chronic timeframes, representing the highest-risk phenotype. The PC loadings for both CVD and CHD can be found in Supplemental Files.

The sensitivity analysis comparing the peak and highest positive temperature thresholds using estimated cardiovascular morbidity/mortality rates versus the crude observed cardiovascular morbidity/mortality rates revealed that sociodemographic covariates provide a significant influence in estimated heat-related excess cardiovascular morbidity/mortality at given thresholds. Additionally, adjusting for sociodemographic factors accounted for an average 1-3°C difference in peak threshold estimates, varying by temporal scale and CVD subtype.

## DISCUSSION

Increased temperatures were associated with statistically significant increases in CVD and CHD heat-related excess mortality rates at the annual, monthly, and daily temporal scales across diverse community areas in Chicago from 2011 to 2022. When daily T_max_ reaches 101.3°F, there was an estimated average of 0.0979 excess CHD deaths per 100,000 people per day (95% CI: 0.0919–0.1040), and an average of 0.0262 excess CVD deaths per 100,000 people per day (95% CI: 0.0164–0.0370) at 96.8°F. Temperature followed a J-shaped curve for daily and monthly excess CVD and CHD mortality, but was linearly associated annually, which is in line with the acute non-linear associations found in prior literature. Several sociodemographic factors were significantly associated with heat-related excess cardiovascular mortality in our models. As median income of a community area increased, CVD and CHD mortality decreased, and as median age of a community area increased, so did CVD and CHD mortality, though the effect of age plateaued at around age 40. No significant associations were observed between temperature and MI or stroke mortality, nor was temperature related to heat-related excess cardiovascular ED visits.

While it is generally known that increased temperatures are associated with adverse cardiovascular outcomes, this area of research is relatively nascent, and the relationship is not yet well understood. Most exposure-response studies of this type have been performed in the last 5 years, with research focusing on CVD comprising less than 25% of the literature despite being the leading cause of death in many countries [22]. Most studies have assessed the impact of extreme temperatures on CVD on a daily or monthly temporal scale over an extended longitudinal period and found that heat exposure increases the burden of heat-related excess mortality and, in some cases, morbidity [6, 10, 11, 13, 41]. There is rationale for studying longitudinal trends in CVD as long-term exposures and behaviors influence acute event onset, however it is unclear how the estimated disease burden shifts by changing the window of exposure [42, 43]. A study from 2020 estimated that an average of 5608 annual deaths were due to heat (≈2500 of which can be attributed to CVD) in a subset of US counties [44]. One study estimated that there were between 5958 and 7144 excess CVD deaths associated with extreme heat in the contiguous US during their 10-year study period using monthly exposure-response windows [11]. Another study used a daily exposure-response framework and found a 1.8% increase in daily CVD mortality for every 1°C increase in T_max_ above a predefined regional threshold [45]. While our study is not directly comparable because we estimated heat-related excess cardiovascular mortality using peak temperature thresholds, we did find that the heat-related excess CVD mortality rate within Chicago was significantly related to annual, monthly, and daily temperatures.

The primary focus of this study was to take an inductive approach to develop CVD-specific thresholds for Chicago for various temporal scales. With these absolute thresholds we were able to estimate absolute values for excess cardiovascular morbidity and mortality attributed to heat exposure. This approach is different than much of the prior literature which applies a deductive approach using preset temperature thresholds to provide a risk value relative to another preset threshold [9, 46]. A deductive approach can be useful, but it also can miss critical temperature thresholds that are disease-specific while also providing relative values that may not be applicable to stakeholders. Our study identified specific points along the temperature spectrum most predictive of increased rates of adverse cardiovascular outcomes and applied those thresholds to estimate the corresponding heat-related excess cardiovascular morbidity and mortality at the community area level. Assuming a population of 3 million residents, we found that on days above 101°F, ≈2.6 CHD deaths in Chicago are heat-related, and for months with an average T_max_ of 86°F, ≈28 CVD deaths in Chicago are heat-related. These absolute values provide utility to population health stakeholders for vulnerability indexing and resource allocation.

The secondary focus of our study was to identify distinct community-level vulnerability profiles based on excess mortality linked to heat exposure across multiple temporal scales. The analysis revealed five typologies of Chicago community areas: Stable, Acute, Chronic, Moderate, and Compound vulnerability. These profiles captured variation in how communities experience excess mortality due to short-term (daily) versus long-term (monthly and annual) heat exposure. Notably, we observed slightly divergent spatial patterns when comparing community clusters for CVD versus CHD mortality. Communities classified as Acute or Compound for CHD were often categorized as Stable or Moderate for CVD. This spatial misalignment suggests that different physiological, environmental, or social mechanisms may underlie heat vulnerability across cardiovascular disease subtypes, and that temporally specific risk pathways may be differentially expressed across communities. From a policy perspective, these findings highlight the importance of tailoring public health strategies to match the temporal nature of vulnerability in each cluster. Communities classified as acutely vulnerable may benefit most from rapid-response interventions like heat warning systems, emergency cooling access, and targeted health messaging in advance of days approaching 100°F. In contrast, areas with long-term vulnerability may require sustained investments in upstream interventions such as chronic disease management, neighborhood-level cooling infrastructure, and greening initiatives. Recognizing and addressing the differing temporal profiles of vulnerability—especially when they diverge by disease subtype—can help ensure that resources are allocated efficiently and equitably, ultimately reducing the burden of heat-related cardiovascular mortality.

The association between heat and specific subtypes of CVD, namely stroke and MI, remains unclear. In studies evaluating the effect of extreme heat on MI incidence, approximately half found a significant association while half did not [47, 48]. Research on stroke outcomes is similarly complicated [49]. Our study found that there were no significant associations between either MI or stroke excess mortality or ED visit rates and increased temperatures. Furthermore, our study found no evidence of increases of heat-related excess cardiovascular ED visits due to temperature, which is in line with the existing literature [41, 50, 51]. Despite plausible biological mechanisms, there is inconsistent evidence that heat and cardiovascular morbidity are related. However, we did find spatial patterns that seemed to suggest that the community areas with the highest heat-related excess CVD mortality rate usually did not overlap with community areas with the highest heat-related excess CVD ED visit rate. This indicates that there may be underlying features not captured by our models that differentiate these two cardiovascular outcomes.

A limitation of this study is the assumption that patient or decedent residential address accurately depicts the heat exposure for a given health event. Although it is possible that the home address at the time of event is not representative of the burden of heat exposure, we believe that it serves as the best method for geocoding the exposome despite its potential shortfalls. Likewise, if a patient or decedent did not have an address, they could not be geocoded, which potentially excluded some of the populations most vulnerable to heat. Another limitation is the representativeness of the ED data. CAPriCORN is the most comprehensive data-sharing network of medical institutions in Chicago, however it does not encompass all healthcare sites in the city. This underrepresentation likely contributes to a bias toward the null for ED visits, particularly on the South and far North sides of Chicago. Though statistical methods to improve representation were considered, we determined that imputation methods involved assumptions that were too strong. This suggests that the true impact of temperature on ED visits may be stronger than what was observed in our results. Future work should seek to understand some of the underlying clinical drivers of cardiovascular morbidity by developing robust computational phenotypes of cardiovascular disease using longitudinal electronic health records. Further work can also incorporate structural elements such a canopy coverage and street density to improve model performance, and investigate other exposure metrics such as mean temperature, minimum temperature, or heat index.

In conclusion, this study provides a novel approach to quantifying heat-related cardiovascular risk by integrating heat-related excess morbidity and mortality estimates across daily, monthly, and annual temporal scales while accounting for both short-term and cumulative heat exposure. By clustering community areas based on shared vulnerability profiles, our method offers a more granular understanding of how extreme heat impacts cardiovascular health during short- and long-term heat exposure. This framework can be replicated in other urban settings to identify high-risk communities and inform targeted interventions. The identification of sociodemographic risk factors, such as education level, income, and racial/ethnic composition, underscores the importance of addressing social determinants of health in climate adaptation efforts. These results can help guide heat early warning systems, urban planning decisions, and resource allocation to reduce the burden of heat-related cardiovascular mortality. As extreme heat events become more frequent and severe due to climate change, refining risk assessment methods and improving public health interventions will be essential in protecting vulnerable populations.

## Data Availability

While patient and decedent data cannot be shared due to HIPAA compliance, all code will be made available.

https://github.com/petergraffy/heat-thresholds

